# Modelling the laminar connectome of the human brain

**DOI:** 10.1101/2021.03.17.21253439

**Authors:** Ittai Shamir, Omri Tomer, Ronnie Krupnik, Yaniv Assaf

**Author notes:** Corresponding author: Ittai Shamir.

## Abstract

The human connectome is the complete structural description of the network of connections and elements that form the ‘wiring diagram’ of the brain. Due to the current scarcity of information regarding laminar end points of white matter tracts inside cortical grey matter, tractography remains focused on cortical partitioning into regions, while ignoring radial partitioning into laminar components. To overcome this biased representation of the cortex as a single homogenous unit, we use a recent data-derived model of cortical laminar connectivity, which has been further explored and corroborated in the macaque brain by comparison to published studies. The model integrates multimodal MRI imaging datasets of both white matter connectivity and grey matter laminar composition into a laminar-level connectome. In this study we model the laminar connectome of healthy human brains (N=30) and explore them via a set of neurobiologically meaningful complex network measures. Our analysis demonstrates a subdivision of network hubs that appear in the standard connectome into each individual component of the laminar connectome, giving a fresh look into the role of laminar components in cortical connectivity and offering new prospects in the fields of both structural and functional connectivity.

## Introduction

The human connectome is the comprehensive structural description of the network of connections and elements that form the human brain’s ‘wiring diagram’. The importance of exploring the connectome lies not only in uncovering patterns of structural connectivity across the brain, but also in understanding the functional states that emerge from it. For these reasons and more, uncovering the complete human connectome has become a crucial and central mission in neuroscientific research (Sporns et al. 2005).

Since its foundation, the Human Connectome Project has lead the mission to map the connectome using non-invasive neuroimaging techniques, showcasing great progress particularly in MRI imaging of white matter connectivity (Van Essen et al. 2013; Setsompop et al. 2013). Concurrently to the progress in white matter imaging through MRI, promising strides have also been made in imaging of the laminar composition of cortical grey matter. The progress in imaging the substructure of cortical grey matter began with establishing that myelination causes shortening of T1 values (Clark et al. 1992). Next, T1 values were linked to myeloarchitecture (Barbier et al. 2002; Duyn et al. 2007; Van Essen et al. 2019), and the correspondence between T1 clusters and cortical layers was established in rat brains (Barazany and Assaf 2012). Subsequent studies have shown that low resolution T1 MRI can also be used to provide information regarding cortical layers in the human brain (Glasser et al. 2014; Shafee et al. 2015; Lifshits et al. 2018). A complete methodological framework was then established for MRI-based quantification and visualization of the cortical laminar composition (Shamir et al. 2019).

Despite growing progress in connectomics, starting from the macroscale and progressing gradually into higher resolutions in meso- and microscales, the field remains inherently limited by the biased representation of the cortex as a single homogenous unit. Due to the current scarcity of information regarding laminar end points of white matter tracts inside the cortical grey matter, tractography remains focused on transverse partitioning of the cortex into regions, while ignoring radial partitioning into laminar components (Jbabdi and Johansen-Berg 2011).

The integration of macrostructural data regarding white matter connectomics and microstructural data regarding grey matter laminar composition poses a promising development in the field of connectomics (Johansen-Berg 2013). A recently published study offers a simplified granularity-based model of cortical laminar connectivity based on published tract tracing and histological findings, with the purpose of integrating white and grey matter datasets derived from multimodal MRI imaging (Shamir and Assaf, 2021a). The model is then further explored in the macaque brain and corroborated in its visual cortex by comparison to published studies (Shamir and Assaf 2021b). The resulting micro-level connectome offers a more detailed and unbiased representation of the connectome and offers new prospects in the field of both structural and functional connectivity on the laminar level.

In this study we model the laminar connectome of the healthy human brain (N=30 healthy subjects) and explore it via a set of neurobiologically meaningful complex network measures. We focus on investigating network hubs in the standard cortical connectome versus those in the cortical laminar connectome.

## Methods and materials

The framework for modelling cortical laminar connectivity (Shamir and Assaf 2021a) was applied on a set of (N=30) healthy human subjects.

### Subjects

Thirty healthy human subjects (N=30), including 14 male and 16 female, 18-78 years old. Subjects were neurologically and radiologically healthy with no history of neurological diseases. The imaging protocol was approved by the institutional review boards of Sheba Medical Center and Tel Aviv University, where the MRI investigations were performed. All subjects signed informed consent before enrollment in the study.

Each subject was scanned on a 3T Magnetom Siemens Prisma (Siemens, Erlangen, Germany) scanner with a 64-channel RF coil. The scans include the following sequences:

1. A standard diffusion-weighted imaging (DWI) sequence, with the following parameters: Δ/d=60/15.5 ms, b max=5000 (0 250 1000 3000 & 5000) s/mm^2^, with 87 gradient directions, FoV 204 mm, maxG= 7.2, TR=5200 ms, TE=118 ms, 1.5×1.5×1.5 mm^3^, 128×128×94 voxels. This sequence was used for mapping the cortical connectome.
2. An MPRAGE sequence, with the following parameters: TR/TE = 1750/2.6 ms, TI = 900 ms, 1×1×1 mm^3^, 224×224×160 voxels, each voxel fitted with a single T1 value. This sequence was used for delineating the cortical surfaces.
3. An inversion recovery echo planar imaging (IR EPI) sequence, with the following parameters: TR/TE = 10,000/30 ms and 60 inversion times spread between 50 ms up to 3,000 ms, 3×3×3 mm^3^, 68×68×42 voxels, each voxel fitted with up to 7 T1 values (Lifshits et al. 2018). This sequence used for characterizing the cortical layers.

### Image processing

The following datasets were analyzed across Brainnetome atlas regions, a connectivity-based parcellation which consists of 210 cortical regions and 36 subcortical subregions (Fan et al. 2016):

1. **Global white matter connectivity analysis** Each DWI dataset was analyzed for global white matter connectivity using global tractography. Traditional tractography involves streamline estimation by inferring connectivity from local orientation fields. It has been proven that streamline estimation experiences difficulties in reconstructing long tracts, due to high false positive rates associated with strong tracts, alongside difficulties reconstructing complex geometries, due to the partial volume effect associated with crossing fibers (Maier-Hein et al. 2017). Global tractography differs from streamline tracking conceptually and methodologically by finding the full track configuration that best explains the measured DWI data. As a result, this tractography method is less sensitive to noise, and the density of the resulting tractogram is more directly related to the data. The analysis was conducted using MRtrix3 software package, which uses a multi-tissue spherical convolution model that accounts for partial volumes (Tournier et al. 2019), similarly to the analysis by Krupnik et al. (2021). For a visualization of the resulting connectivity matrix of the 30-subject average, see figure 1a (below).

**Fig. 1.**
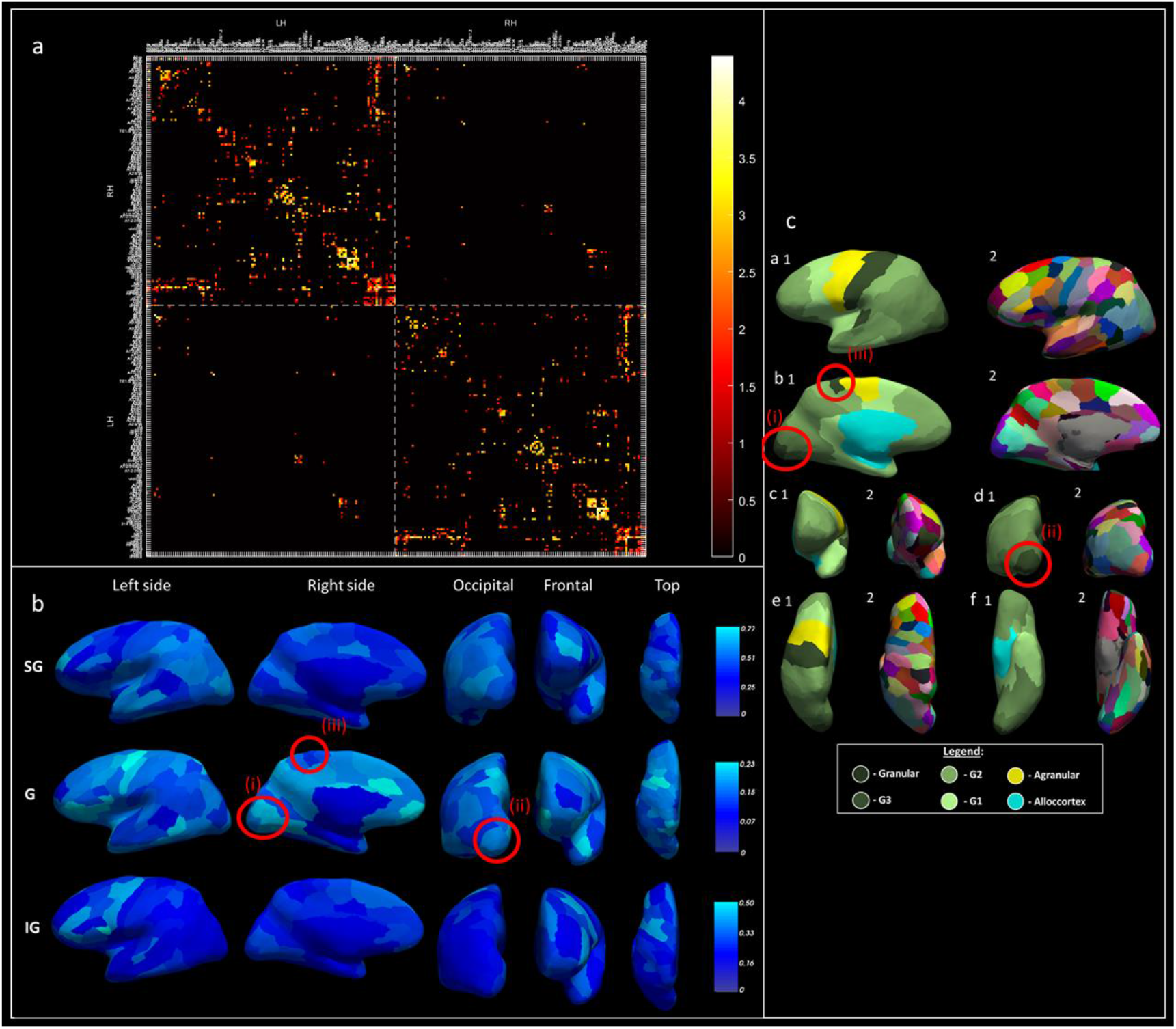
Average global white and gray matter datasets and estimated granularity indices across Brainnetome atlas regions: a- Average cortical connectivity (of all 30 subjects) across left (LH) and right (RH) hemispheres: average connectivity matrix, representing (log(number of tracts)) for connections that appear in at least 75% of subjects (color scheme adapted from Charles 2021) b- Average cortical laminar composition (of all 30 subjects) across left hemisphere: where: top row-supragranular layers (SG), middle row-granular layer (G), bottom row-infragranular layers (IG), and columns represent different viewpoints c- Granularity indices (left- 1) and Brainnetome atlas regions (right- 2) across left hemisphere, from several viewpoints: left side (a), right side (B), front (C), occipital (D), top (E), and bottom (F) Features in b and their respective counterparts in c correspond to unique granular presence (circled in red): features (i) and (ii): high presence of granular laminar component in V1 (in b), in correspondence with a high granularity index (in c); feature (iii): low presence of granular laminar component in M1 (in b), in correspondence with a low granularity index (in c)
2. **Cortical laminar composition analysis** Each IR EPI dataset and its corresponding MPRAGE were analyzed for cortical laminar composition using our original framework, according to the following four principal steps:
  a. *IR decay function fit*: the low resolution fast echo planar imaging inversion recovery (EPI IR) protocol was utilized for estimation of multiple T1 components per voxel using the following IR decay function fit (Lifshits et al. 2018):

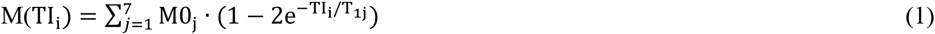

Where: M(TI_i_)- Magnetization at the *i*^*th*^ inversion recovery image, in other words the magnetization measured for each specific T1 component M0_j_- Predicted magnetization at TI=0ms for each T1 component (j) in the voxel T_1j_- Longitudinal relaxation time for each T1 component j was set up to 7 for the low resolution experiments, indicating fit to seven individual exponential fits, based on the assumption that there are 7 T1 components in the tissue – 1 for CSF, 1 for WM and heavily myelinated layer of the cortex and additional 5 cortical layers. Normalization of each of the predicted magnetization values according to 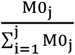 then represents the voxel contribution of each corresponding T1 component (j).
  b. *T1 probabilistic classification:* T1 values were then assigned to different brain tissues and utilized to extract the subvoxel composition of each T1 layer (Lifshits et al. 2018; Barazany and Assaf 2012; Peel et al. 2000; Shamir et al. 2019; Lotan et al. 2021). The T1 classification process involved fitting the T1 histogram to a probabilistic mixture model consisting of t-distributions. The probability of each t-distribution in the voxel was calculated according Bayes’ formula:

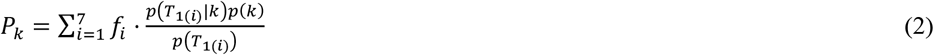

Where: *k*- A specific t-distribution *T*_1(*i*)_- T1-value of the *i*^*th*^ component of the voxel *f*_*i*_- Partial volume of *T*_1(*i*)_ (normalized as show in previous section) *p*(*T*_1_)- General whole-brain probability of a *T*_1_-value *p*(*k*)- Probability of t-distribution k *p*(*T*_1_|*k*)- Probability of the *T*_1_-value in t-distribution *k* Fit to 18 t-distributions was deemed satisfactory according to evaluation of the Bayesian information criterion for 1-20 t-distributions. Each resulting group of distributions then corresponds to various types of brain tissue:
    i. White matter (WM) characterized by low T1 values, represented by t-distribution 1-3.
    ii. Gray matter (GM) characterized by mid-range T1 values, represented by t-distributions 4-9, corresponding to 6 T1 layers. Since T1 is considered a measure of myelination (Clark et al. 1992), T1 layers with higher indices (or smaller T1 values), are more myelinated and are therefore located deeper in the cortical cross section. t-distributions 4,5,6,7,8, and 9 are then termed T1 layers 6,5,4,3,2 and 1 (respectively).
    iii. Cerebral spinal fluid (CSF) characterized by high T1 values, represented by t-distributions 10-18.
  c. *Cortical volume sampling:* We then implemented a geometric solution to cortical sampling based on a system of virtual spheres dispersed throughout the entire cortex. A sphere was chosen as a robust alternative to cortical normals due to its symmetry and invariance to rotation (Shamir et al. 2019). The sampling process started with delineation of the inner, mid, and outer cortical surfaces using the *FreeSurfer* pipeline (Fischl 2012). The virtual spheres were then generated with centers on the mid surface and edges tangential to both the inner and outer cortical surfaces. Each hemispheric volume consists of ∼75,000 spherical volumes with and average radius of ∼1 mm.
  d. *Data sampling:* To sample the high resolution spheres in the low resolution T1 dataset (3^3^ *mm*^3^), a super-resolution solution was implemented. The solution involved partitioning each voxel into 10^3^ subvoxels, each assigned location properties, primarily their location inside or outside of a given sphere. Spherical volume weights were then assigned to each sphere, corresponding to each voxel’s contribution to its spherical volume, according to the following:

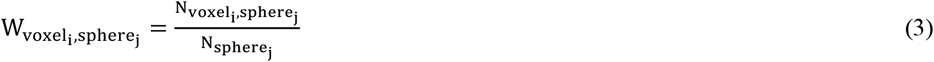

Where: 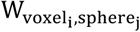- Volume weight of voxel i per sphere j 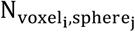- Number of subvoxels from voxel i located inside sphere j 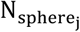- Total number of subvoxels located inside sphere j The cortical composition of each sphere was then estimated by multiplying the volume weights of each sphere by their corresponding voxel probability maps (see *T1 probabilistic classification*). The process was repeated across all spheres, according to the following equation:

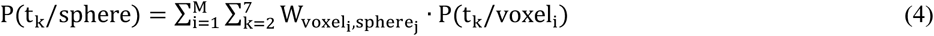

Where: P(t_k_/sphere)-Probability of t-distribution k per sphere k- t-distributions 4,5,..,9, representing T1 layers 6,5,..,1 (respectively) M- Number of voxels within which sphere j lies 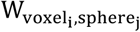 - Volume weight of voxel i per sphere j P(t_k_/voxel_i_)-Probability of t-distribution k in voxel i The resulting whole-brain cortical laminar composition is then simplified by grouping the layers into three laminar components:
    i. Supragranular (SG) laminar component, which includes T1 layers 1, 2, and 3.
    ii. Granular (G) laminar component, which includes granular T1 layer 4.
    iii. Infragranular (IG) component, which includes T1 layers 5 and 6.
    For a visualization of the resulting cortical laminar composition of the three laminar components, averaged across all 30 subjects, see figure 1b (below).
3. **Model of cortical laminar connectivity** The multimodal MRI datasets were integrated using the data-derived, granularity-based model of cortical laminar connectivity (Shamir and Assaf 2021a, b). The model includes a set of whole-brain laminar-level connectivity rules that integrate white matter connectomics on a macroscale and grey matter composition on a mesoscale, using a set of granularity-based connectivity rules. To implement the model rules, each region in the Brainnetome atlas was labelled according to its granularity index (see figure 1c below). The labelling process was conducted manually according a an adapted von Economo-Koskinas atlas (Solari and Stoner 2011; Beul and Hilgetag 2015), and similarly to the labelling by Shamir and Assaf (2021a). Each Brainnetome atlas region then holds three components, corresponding to its white matter connections, its grey matter laminar composition, and its granularity index. These components are crucial for the model application, since its rules use weighting of white matter tracts according to the cortical laminar compositions of the connecting regions based on their respective granularity indices. The model then addresses two types of cortical connections:
  a. *Tractography-based (long-range) connections:* tractography-based connections represent most of the connections in the model, including relatively stronger connections between different cortical regions, as well as connections between the cortex and the subcortex. They are expanded to the laminar level according to the rule of connectivity that corresponds to the connecting regions, their granularity indices, and their laminar compositions.
  b. *Assumed (short-range) connections:* assumed connections include vertically-oriented connections between different laminar components within a single cortical region. They are modelled as relatively weaker connections in the model according to the laminar composition of each cortical region.
  For a comprehensive description of the model of cortical laminar connectivity and its data-derived origin, see (Shamir and Assaf 2021a). The resulting laminar connectome includes three times as many nodes as the standard connectome since each original regional node now consists of three laminar locations in that same specific region. A full visualization of the 30-subject average of the resulting model of laminar connectivity can be seen in figure 2 parts a-b (below). The source code of the average standard and laminar connectomes, as well as the complete code for modelling a laminar-level connectome, are freely available for noncommercial use (at github.com/ittais/Laminar_Connectivity).

**Fig. 2.**
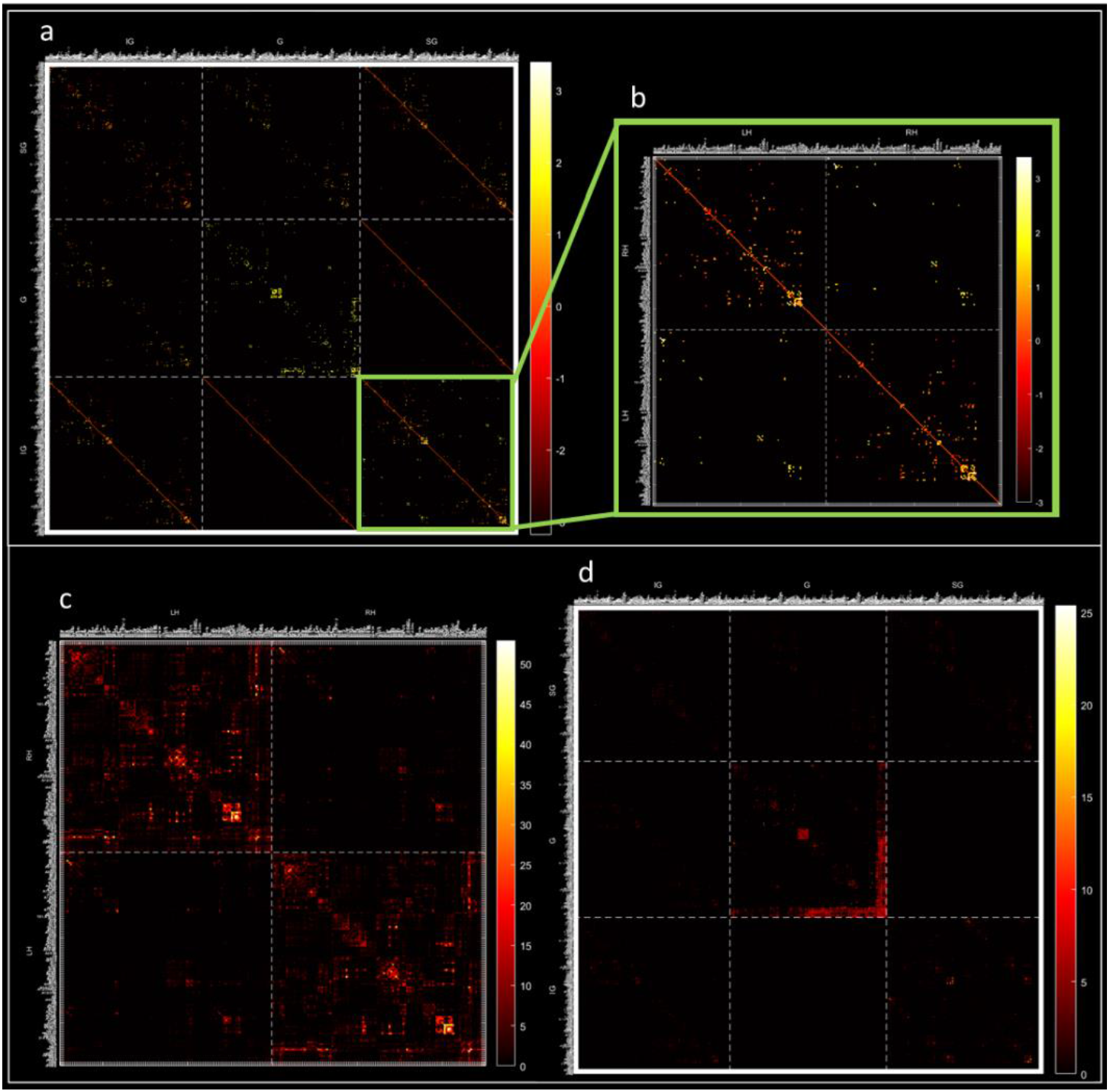
30-subject average cortical laminar connectivity (top) and standard deviations for both connectomes (bottom) across Brainnetome atlas regions: a- Average supra-adjacency matrix, representing whole-brain laminar-level connections, where the following abbreviations correspond to laminar components: IG- infragranular, G- granular, SG- supragranular. Model results are displayed as log(number of tracts) for all connections that appear in at least 75% of subjects b- A closer look at the supragranular-supragranular component of the average supra-adjacency matrix c- Standard deviation of standard cortical connectomes d- Standard deviation of cortical laminar connectomes (color schemes adapted from Charles 2021) The variability of the resulting connectomes, representing both standard connectivity and laminar connectivity across subjects (N=30), are then evaluated (see figure 2 parts b-c). Subsequently, both connectomes are then explored via a set of neurobiologically meaningful complex network measures (Rubinov and Sporns 2010).
4. **Complex network analysis** Once the standard connectome was extracted and the laminar connectome was modelled, we used tools for network analysis to explore their connectivity patterns and unique network features:
  a. **Network complexity:** both connectomes were initially tested for non-trivial topological features that occur in complex networks, such as a heavy tail in degree distribution, but do not occur in simple networks, such as random graphs (see figure 3 below).

**Fig. 3.**
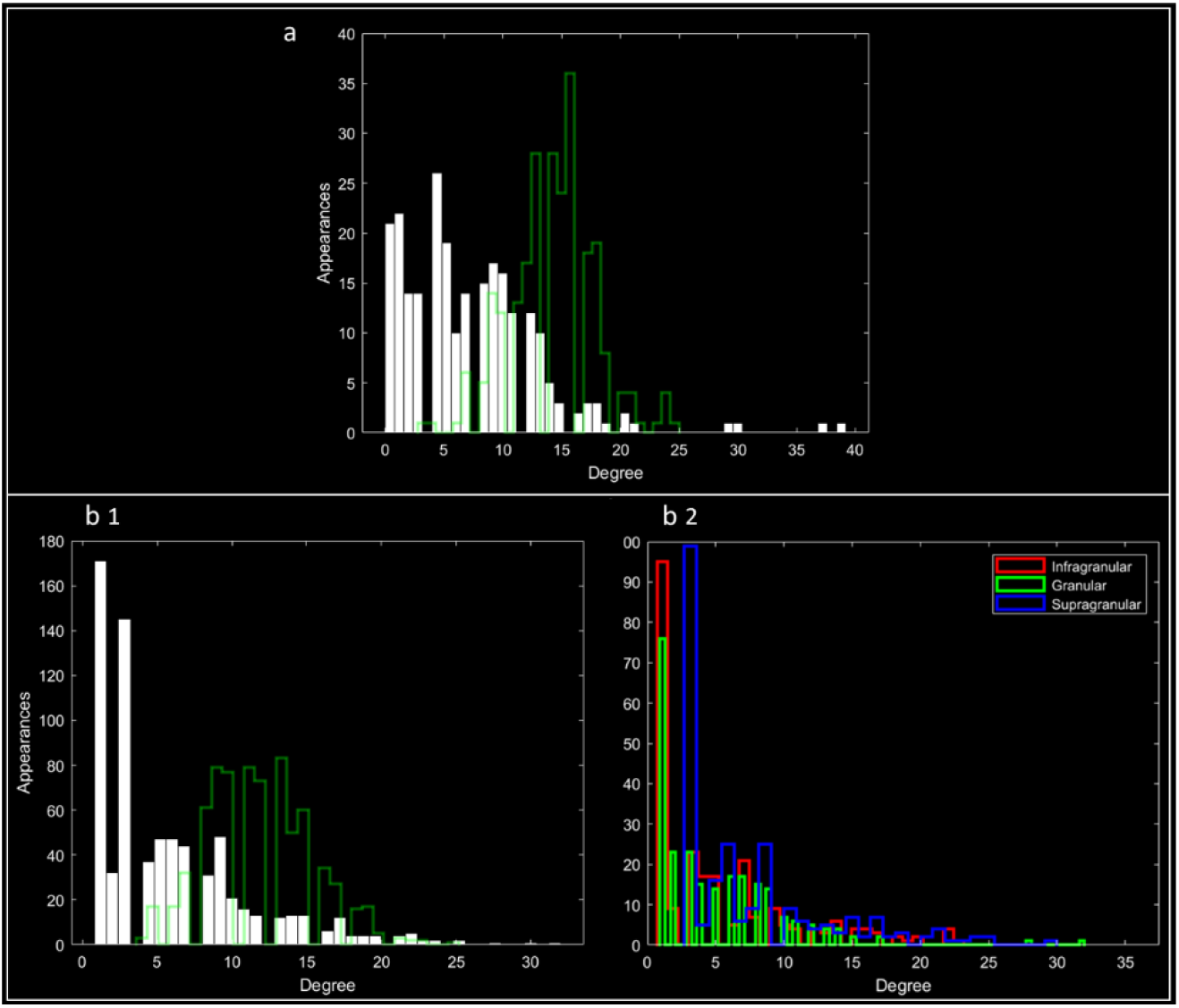
Degree distributions: Distributions of degrees for both the standard cortical connectome (a), as well as the cortical laminar connectome (b1), of the average connectomes for all connections that appear in at least 75% of subjects. Each of the two is presented against a random network with an equal number of nodes and edges (histogram outlines in green). For the laminar connectome, distributions for each laminar connectome are colored individually (b2): infragranular (IG)- red, granular (G)- green, supragranular (SG)- blue Notice the positive skew, or “heavy-tail”, in degree distributions in both cases and expressly in the laminar connectome
  b. **Network analysis:** once the complexity of both networks was shown, we explored and compared the average standard connectome to the components of the average laminar connectome. Global connectivity measures, including global efficiency and connectivity density, were chosen for evaluating the cost-efficiency trade-off. Additionally, local connectivity measures of centrality were calculated, including node degree and strength.

For a schematic representation of our methodology for modelling and analyzing the laminar connectome, see Supplementary Material figure 1.

## Results

After calculating average matrices for both standard cortical and cortical laminar connectomes, the variability of both connectomes was evaluated across all N=30 subjects (see figure 2 parts c-d). The standard connectome exhibits relatively higher standard deviation values, that correspond to the overall higher range of connectivity values. The subcorticocortical connections exhibit relatively higher standard deviation within each matrix, specifically in the granular to granular connections for the laminar connectome. This can be attributed to the higher connection values of subcorticocortical connections, relative to corticocortical connections.

Two measures of global network connectivity were calculated for both the average standard and average laminar connectomes:

1. Global efficiency- an efficiency measure representing the average inverse shortest path length in the network.
2. Connection density- a cost measure representing the number of edges in a network as a proportion of the maximum possible number of edges.

The two measures where then plotted against one another to evaluate the cost-efficiency trade-off (Bullmore and Sporns 2012) in both connectomes (see figure 4).

**Fig. 4.**
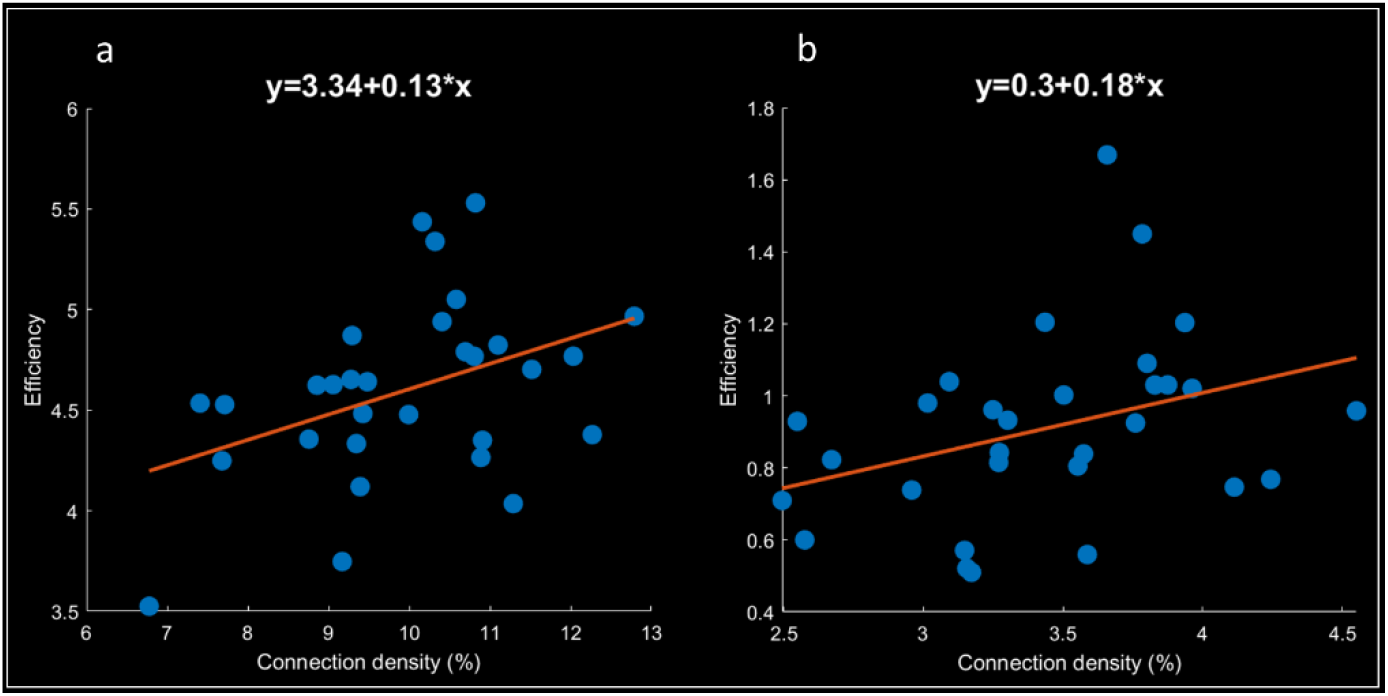
Cost-efficiency trade-offs of both connectomes: Global efficiency and connection density values, fitted to a linear regression model (top), for all N=30 subjects, including the standard cortical connectomes (a) and the cortical laminar connectomes (b)

Examination of the cost-efficiency trade-off in the cortical laminar connectome shows that both the connection density as well as the global efficiency are reduced compared to the standard connectome. Connection density is a value inversely related to the maximum possible number of edges in the standard connectome: 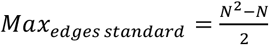, where N is the number of nodes in the standard connectome network. The number of nodes in the laminar connectome is 3 * *N*, since the cortex is divided into three cortical components. Accordingly, the maximum possible number of edges in the laminar connectome for big networks (large enough N): *Max*_*edges laminar*_ ∼9 * *Max*_*edges standard*_. Since the density is inversely related to the maximum possible number of edges, theoretically the ratio should be: 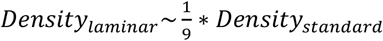. In practice, the laminar connectome exhibits higher density than expected that instead follows: 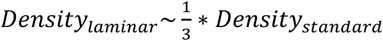 (see density value range in figure 4). Global efficiency is a value inversely related to the mean shortest path (MSP), where: *MSP*_*standard*_ *αN*. Since the laminar connectome has 3 * *N* nodes, the efficiency of the laminar connectome should follow: 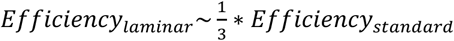. In this case, the laminar connectome does roughly follow this ratio (see efficiency range in figure 4). Consequently, the efficiency to density ratio for the laminar connectome is slightly higher than for the standard connectome.

To examine the connectivity patterns of both the standard connectome as well as the cortical laminar connectome, we conducted an analysis of a two neurobiologically meaningful network measures (Rubinov and Sporns 2010). The following two measures of centrality were included:

1. Degree- the number of edges connected to a node (see figure 5, below).

**Fig. 5.**
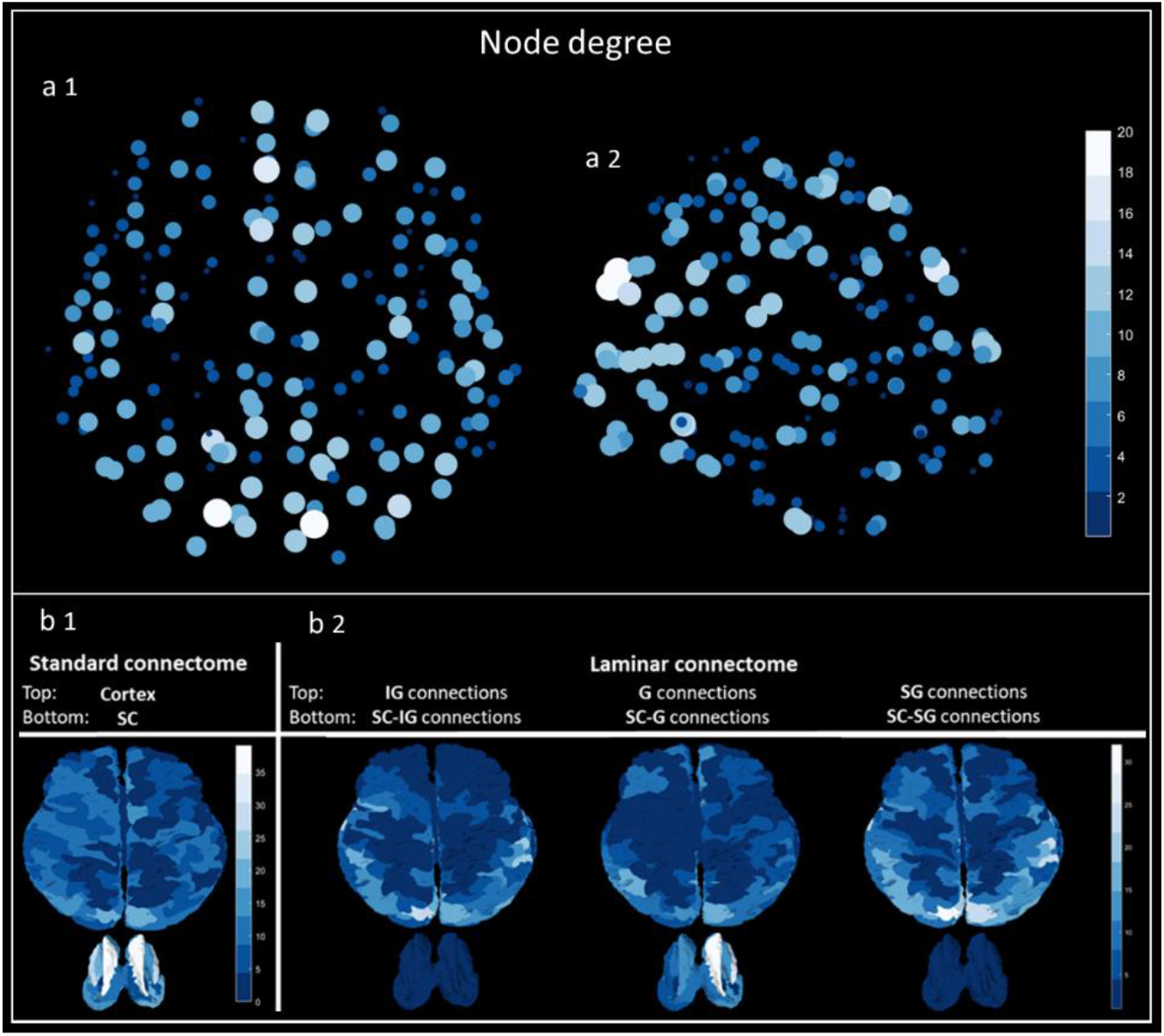
Node degree: a- Distribution of degree values for the average standard connectome across cortical regions of the Brainnetome atlas, top view (a1) and lateral view (a2) b- Whole-brain distributions of degree values for the average standard connectome (left column) compared to the average laminar connectome, including: infragranular (IG), granular (G) and supragranular (SG) components (3 right columns, left to right). Each connectome depicts cortical connections (top) and subcortical connections (bottom). For each laminar component, cortical connections include connections between the specified component and all other components (top), and subcortical connections include connections between the specified component and the subcortex (bottom)
2. Strength- sum of all neighboring edge weights (see figure 6, below)

**Fig. 6.**
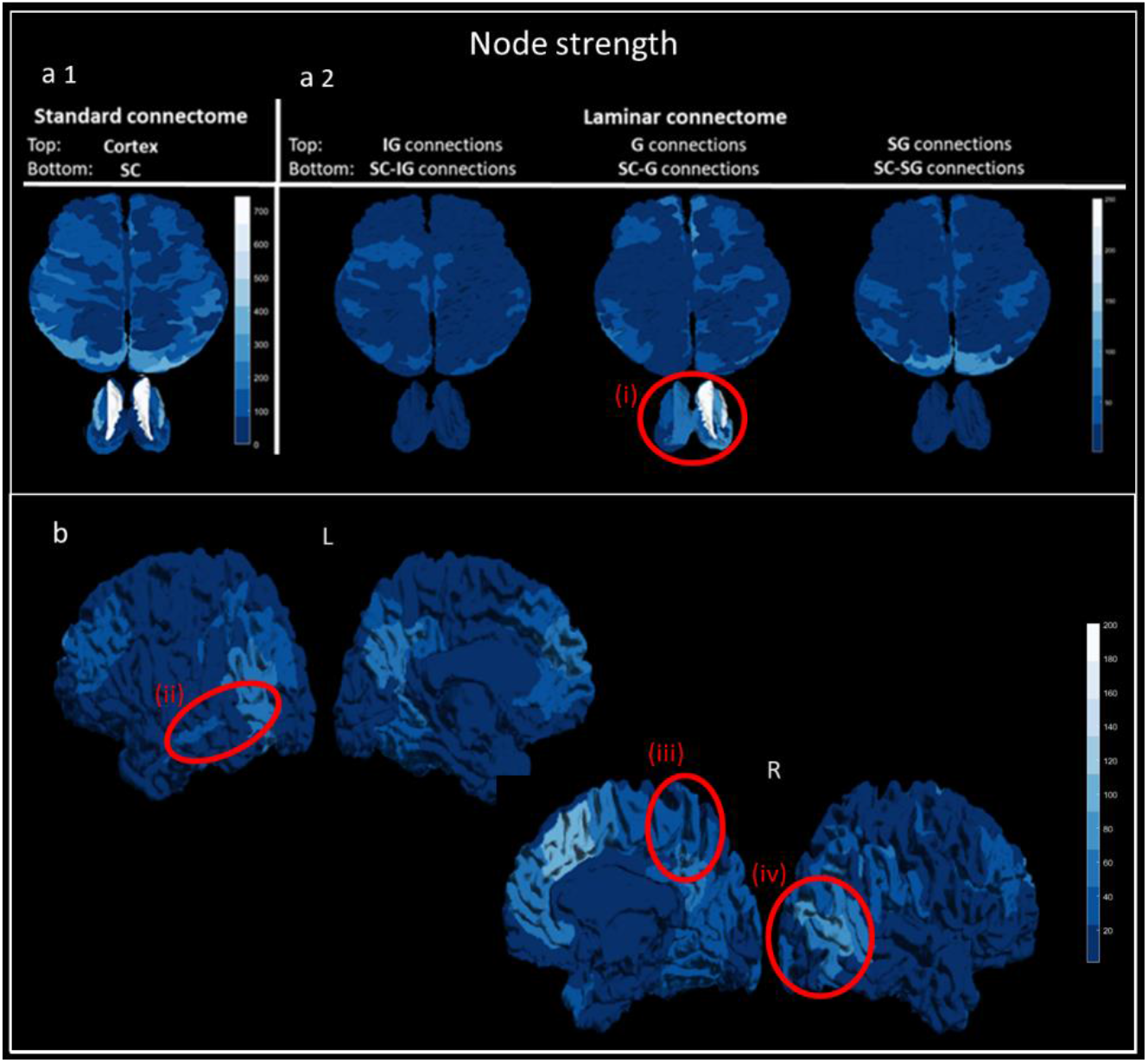
Node strength: a- Whole-brain distributions of strength values for the average standard connectome (left column) compared to the average laminar connectome, including: infragranular (IG), granular (G) and supragranular (SG) components (3 right columns, left to right). Each connectome depicts cortical connections (top) and subcortical connections (bottom). For each laminar component, cortical connections include connections between the specified component and all other components (top), and subcortical connections include connections between the specified component and the subcortex (bottom). Feature (i) (circled in red) shows high centrality of subcortical regions for the granular component of the laminar connectome b- Distribution of strength values for the granular (G) component of the laminar connectome across both left (L, top) and right (R, bottom) hemispheres, from lateral view (left) and sagittal view (right). Features (circled in red) show high strength values in the auditory cortex (ii), as well as the primary motor (iii) and primary visual (iv) cortices

For an exploration of additional measures, including clustering coefficient, local efficiency, and core/periphery, see Supplementary Material figure 2.

When examining the degree values across cortical regions of the standard connectome, a well-established pattern of node centrality appears in the occipital, temporal, and medial parietal cortices (Sporns 2009). When examining the degrees across both connectomes, subcortical regions (thalamus, caudate, putamen and hippocampus) alongside a set of cortical regions appear as hubs with high degree values, including: superior frontal regions (A9m, A10m), superior parietal regions (A7 subregions), superior occipital regions (msOccG) and the cuneus (rCunG). These hubs exhibit both high degree and high strength in the standard connectome, as well as cumulatively across components of the laminar connectome. The infragranular and supragranular components do not include the subcortex but include cortical hubs, while the granular component includes the subcortex and mainly non-frontal hubs such superior parietal and superior occipital regions.

When examining the strength (weighted degree) values, a similar more generalized pattern appears. In the standard connectome, the subcortex overpowers the distribution across nodes and exhibits the highest strength values, followed by occipital and parietal regions. In the laminar connectome, the granular component exhibits high strength values in the subcortex. A closer examination of the granular component demonstrates high strength values in the auditory cortex, alongside the primary motor and primary visual cortices.

## Discussion

In this study we model and explore the laminar connectome of N=30 healthy subjects using multimodal MRI imaging datasets. These datasets include both white matter connectivity and grey matter laminar composition, which are integrated using our novel model of cortical laminar connectivity (Shamir and Assaf 2021a, b). The resulting laminar-level connectome is then explored across the Brainnetome atlas regions (Fan et al. 2016) using a set of neurobiologically meaningful complex network measures (Rubinov and Sporns 2010), while focusing on comparison of the cortical laminar connectome to standard cortical connectome.

When we examine both connectomes on a global scale, several similarities and other differences appear. On the similarity side, both connectomes exhibit a heavy tail in their degree distributions, which is even more noticeable in the laminar connectome. A heavy tail in a degree distribution is a non-trivial topological feature that occurs in complex networks but does not occur in simple or random networks. This feature is also associated with higher network resilience to nodal removal (regional damage). On the difference side, the standard connectome exhibits relatively higher standard deviation values across subjects, relative to the laminar connectome. This higher variability corresponds to the overall higher range of connectivity values in the standard connectome. Nonetheless, both connectomes exhibit higher variability in subcorticocortical connections, which can once again be attributed to stronger connections relative to corticocortical connections. An additional difference between the two connectomes appears when exploring the cost-efficiency trade-offs, by plotting global efficiency against connection density. The laminar connectome exhibits higher connection density than expected based on the relative number of nodes in the network alone. With regards to efficiency, the laminar connectome exhibits a relatively lower global efficiency, as expected based on number of nodes alone. However, the efficiency to density ratio is maintained and even slightly elevated for the laminar connectome in comparison to the standard connectome.

When we examine both connectomes on a local scale using complex network measures, we get a more nuanced image of connectivity patterns across the brain. Several features appear in the granular component of the laminar connectome, including high subcortical centrality on a whole-brain level, alongside high regional centrality in the auditory cortex, M1, and V1 on a cortical level. In addition, the distributions of degree and strength values in the standard connectome reestablish the notion of a rich-club bihemispheric organization that includes subcortical regions (van den Heuvel and Sporns 2011, 2013). The distribution of these values across components of the laminar connectome exhibits a cumulative nature, which is strengthened upon further examination of the core/periphery partitioning in the standard connectome (for an examination of inter-subject consistency, see Supplementary Material figure 3). The standard connectome displays a core of highly interconnected hubs across the cortex and subcortex, a subset of these hubs appears in each individual component of the laminar connectome. The infragranular component displays hubs across non-frontal cortical regions, the granular component displays hubs that include subcortical regions, and the supragranular component displays hubs that include frontal cortical regions.

This study presents an innovative exploration of a network model of the healthy human connectome on the laminar level. This model embodies a simplified set of laminar-level rules of connections, based on a systematic review of histological tract tracing studies (Shamir and Assaf 2021a). While this model has been corroborated ex-vivo in the macaque brain (Felleman and Van Essen 1991; Shamir and Assaf 2021b), corroborating the results in-vivo in the human brain is a more nuanced task. However, our network analysis showcases several expected features, mainly concerning high centrality of granular connections to visual, motor, and auditory regions.

The characterization of the healthy human laminar connectome presented here could support the investigating of pathologies that are assumed to involve abnormalities in layer-dependent cortical connectivity, such as schizophrenia and autism. Using the network modelling and analysis framework presented here, the patterns of connectivity behind such conditions could be further explored and elucidated.

## Data Availability

Data will be available upon request.

## Statements & Declarations

### Funding

The authors declare that no funds, grants, or other support were received during the preparation of this manuscript.

### Competing Interests

The authors have no relevant financial or non-financial interests to disclose.

### Data Availability

The datasets generated during and/or analyzed during the current study are available in the following GitHub repository: https://github.com/ittais/Laminar_Connectivity

### Ethics approval

This study was performed in line with the principles of the Declaration of Helsinki. The imaging protocol was approved by the institutional review boards of Sheba Medical Center and Tel Aviv University, where the MRI investigations were performed.

### Consent to participate

Written Informed consent was obtained from all individual participants included in the study.

### Consent to publish

The authors affirm that human research participants provided informed consent before enrollment in the study, including consent to publish the images resulting from the study.

## Notes

### Competing Interest Statement

The authors have declared no competing interest.

### Funding Statement

No funding to disclose.

