## Supplementary Material for "Modelling the laminar connectome of the human brain"

Ittai Shamir [a], Omri Tomer [b], Ronnie Krupnik [b], Yaniv Assaf [a,b]

[a] Department of Neurobiology, Faculty of Life Sciences, Tel Aviv University, Tel Aviv, Israel

[b] Sagol School of Neuroscience, Tel Aviv University, Tel Aviv, Israel

### Methods and materials

For a full schematic visualization of our streamline process for modelling and analyzing the cortical laminar connectome, see figure 1 (below).

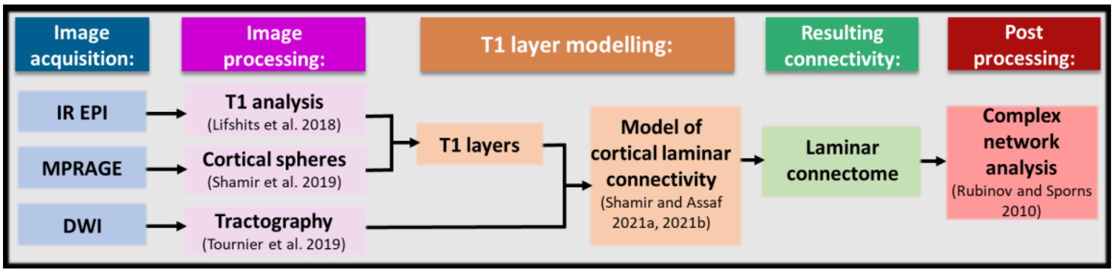


**Fig. 1** A schematic representation of our methodology for modelling and analyzing the laminar connectome (left to right)

### Results

To examine the connectivity patterns of both the standard connectome as well as the cortical laminar connectome, we conducted an analysis of a several additional neurobiologically meaningful network measures (Rubinov and Sporns 2010). The additional measures include the following:

1. Clustering coefficient- the fraction of a node's neighbors that are also neighbors of each other
2. Local efficiency- the global efficiency (the average inverse shortest path length in the network) computed on node neighborhoods
3. Core/periphery- a partition of the network into two non-overlapping groups of nodes (core and periphery), maximizing within core edges and minimizing within periphery edges.

For an exploration of the above-mentioned network measures, see figure 2 (below).

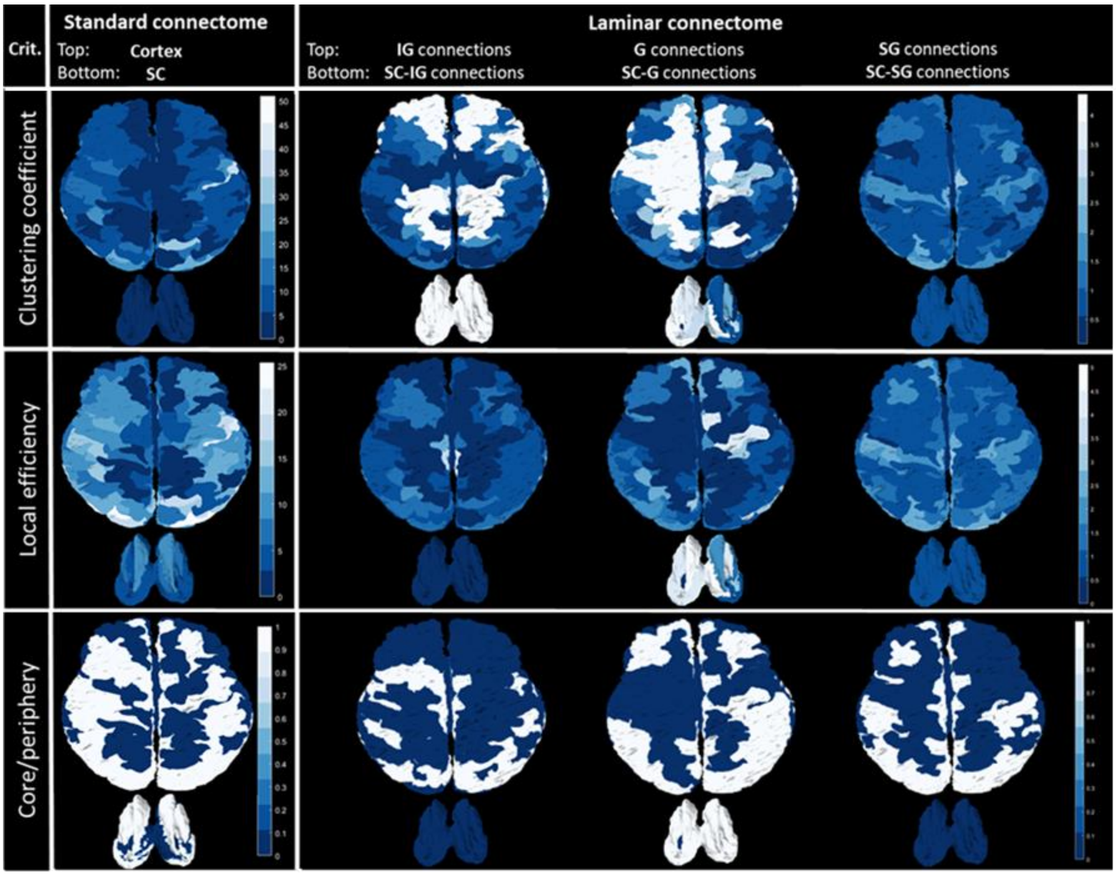


**Fig. 2** Comparison of average connectomes:

The average standard connectome (left column) compared to the average laminar connectome, including: infragranular (IG), granular (G) and supragranular (SG) components (3 right columns, left to right). Each connectome depicts cortical connections (top) and subcortical connections (bottom). For each laminar component, cortical connections include connections between the specified component and all other components (top), and subcortical connections include connections between the specified component and the subcortex (bottom). The rows depict network measures, including (from top to bottom): clustering coefficient, local efficiency, and core/periphery

Clustering coefficient is a measure of network segregation, which represents specialized processing in clusters of nodes, and local efficiency is a measure of network integration, which represents the ability of the network to combine information. When examining both measures in the standard connectome, temporal (A37), occipital (mOccG) and frontal (A44) regions exhibit high clustering and efficiency values. In comparison, across the laminar connectome the infra- and supragranular components exhibit a more even distribution across cortical regions, while the granular component exhibits high clustering and efficiency values in subcortical regions.

With regards to dissimilarities between the measures, it appears that in the standard connectome the subcortex exhibits a low clustering coefficient and high local efficiency. The distribution of local efficiency values across the standard connectomes is more even, compared to higher clustering coefficient values specifically in frontal regions. Lastly, the network hubs are more pronounced for clustering coefficients in the laminar components, and more pronounced for local efficiency in the standard connectome. Nonetheless, several similar patterns appear across these two measures, primarily hemispheric symmetry in values and high values for subcortical regions in the supragranular component of the laminar connectome. Additionally, these measures exhibit several hubs in occipitotemporal regions.

For an examination of inter-subject consistency of core/periphery partitioning in both the standard connectome and the laminar connectome, see figure 6-1 (below).

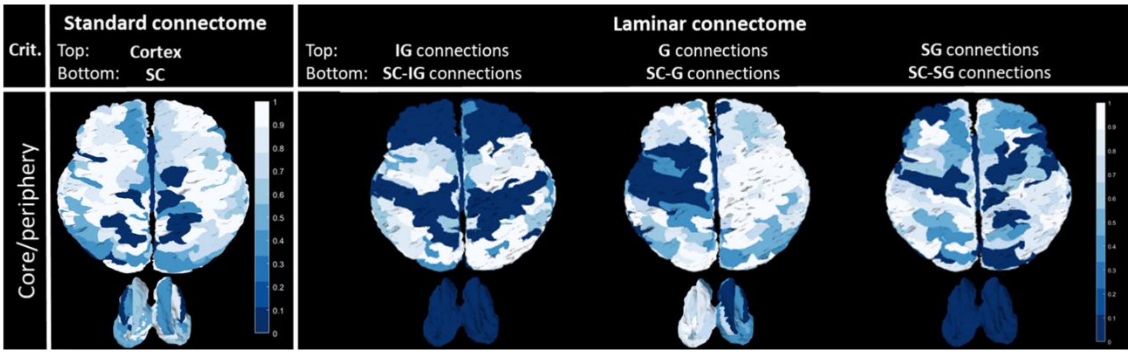


**Fig. 3** Inter-subject consistency of core/periphery values:

Standard deviation of core/periphery partitioning across subjects (N=30 subjects) for both the standard connectome (left column) and the laminar connectome, including: infragranular (IG), granular (G) and supragranular (SG) components (3 right columns, left to right). Each connectome depicts cortical connections (top) and subcortical connections (bottom). For each laminar component, cortical connections include connections between the specified component and all other components (top), and subcortical connections include connections between the specified component and the subcortex (bottom)

When examining the core/periphery in both the standard and laminar average connectomes, less hemispheric asymmetry is exhibited, particularly in components of the laminar connectome (as opposed to the high hemispheric symmetry exhibited in both degree and strength values).
